# Setting Limits on Neural Network’s Predictive Capacity in T1D Blood Glucose Concentration

**DOI:** 10.1101/2020.08.04.20117812

**Authors:** Thomas Chuna

## Abstract

The ability to anticipate changes in blood glucose (BG) concentration would have a great impact on Type 1 diabetics (T1D). In order to create T1D treatment plans, patients collect a BG concentration time series. It has been demonstrated that various types of recurrent neural networks, such as Long Short Term Memory (LSTM), have success forecasting T1D BG concentrations. However, limited work has been done to characterize the T1D time series or set limits on neural network’s predictive capacity. In this work, a T1D patient’s 14 day BG concentration time series is studied. First, I test the time series stationarity. Then I use auto-correlation analysis, spectral analysis, and Gaussian process regression to characterize the T1D BG time series. Finally, the LSTM’s prediction quality is quantified and interpreted at different prediction intervals. The success or failure of the LSTM’s predictions are interpreted using the characterization of the time series.

## 1 Introduction

An overview of Diabetes is provided in App. A. Medical professionals require T1D patients to track their blood sugars, exercises, medication, and meals so that their treatment plans can be adjusted. Type one diabetic (**T1D**) patients are creating a time series by recording their blood glucose (**BG**) concentrations. I note my supply of data came directly from a T1D patient. They have access to the data generated by their continuous glucose monitor (**CGM**) records. There is a 5 min sampling rate yielding approximately 288 samples per day. Giving more than 4000 BG measurements in this data set. A review of glucose monitoring methods is provided in App. B. I provide a visualization of the patient’s CGM BG records in Fig.1. The BG measurements can be understood as samples from a *stochastic process, y*(*t*). Time series analysis (*i*.*e*. the analysis of stochastic processes), is discussed in App. C.

**Figure 1:**
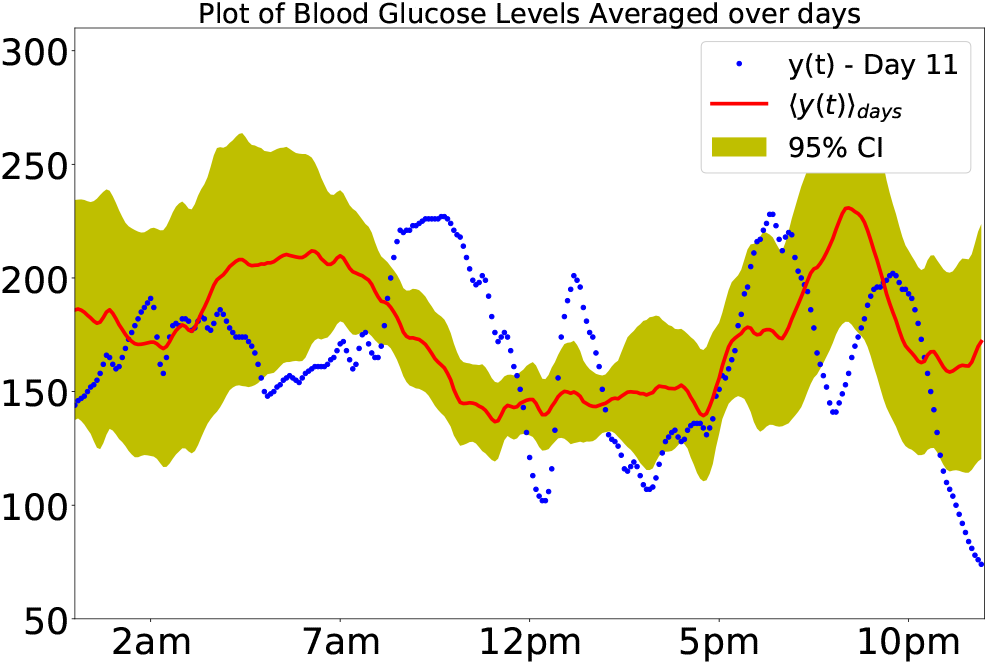
Plot of time series average over the time of day alongside the blood glucose concentrations of day 11 for comparison.

This study will only focus on CGM records collected over 14 days from a real T1D patient. ^1^ In this work, Long-Short term memory (**LSTM**) neural network is trained to predict the BG levels of a patient given their prior blood sugars. Similar work has been done in [1, 2, 3, 4, 5]. I refer the reader to [1, 3] for descriptions of the LSTM method. An example LSTM is provided by Martinsson https://github.com/johnmartinsson/blood-glucose-prediction. The significance of this work is NOT in demonstrating the success of the LSTM over other methods, this has already been done [3]. Rather, this work assesses the properties of BG data and the limits of LSTM BG predictions.

## 2 Project Methods

First, I examine the stationarity of the time series, (App. C). This done by considering two different averages: average across days, ⟨*y*(*t*)⟩_days_, Fig.1, and average over all measurements, 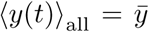. Even though the stationary property is a fundamental to time series analysis, there are no explicit calculations made in the surrounding literature. This seems to be an important first step towards understanding the application of LSTM’s.

Second, I examine the autocorrelation (**ACor**) in the BG time series, (App. C). Stationarity ensures that the Acor can be calculated. For a given time interval, *t*_*p*_, the Acor is the correlation between BG measurement, *y*(*t*_0_) and measurement, *y*(*t*_0_ + *t*_*p*_). The autocorrelation function gives an indication of how long the body remembers its physiological state. The Fourier transform of the Acor function (i.e. spectral density) can give an indication of the complexity/dimensionality of the time series. The spectral analysis of the Acor function will serve as a proxy, supplying similar intuition as dimensional reduction would for AI. Due to the complexity of the underlying physiological model the Fourier transform of the BG time series does not yield useful results.

Third, Since Fourier transform fails, we gain intuition about CGM sampling rates using Gaussian process regression (**GPR**). GPR differs from linear regression by its assumption that collection of *n* BG samples are the coordinates of a random sample from an *n*-dimensional Gaussian (App. D). This enables GPR to produce non-linear estimates. I rarefy the CGM data to produce different sampling rates, then apply GPR to the various rates. I use plots and root mean squared error (**RMSE**) to compare the fits at various sampling rates.

Lastly, I train the LSTM to predict future blood sugars in the time series. I test the LSTM on a portion of the time series completely unseen during training. This testing set estimates the LSTM’s real-world predictive capacity quantitatively via RMSE. It is expected the LSTM predictions to get worse or better depending on the prediction interval *t*_*p*_. I study this dependence and explain it in terms of physiological models and autocorrelation.

## 3 Stationarity of CGM Time Series

### 3.1 Stationarity Across All Samples

For this study, we use a T1D patient’s 14 day CGM times series. It is important to consider that each patient has their own unique average blood sugar (**A1C**) and this A1C is tested every three months. We approximate the A1C as this average over all measurements, regardless of time of day, ⟨*y*(*t*)_all_⟩. As a diabetic, this A1C is different at each doctor visit, the protocol is not to retest A1C within three months of the last measurement. By this very fact, we can know that the time series is NOT *stationary*, except on some small sampling duration. 14 days is a sixth of the three month upperbound. Numerically, I jackknife bin over the CGM data by days, (described in [6]) and calculate the mean and standard error of the jackknife bins Fig. 2. I find that there is no strong statisical evidence against assuming stationarity across all BG samples.

**Figure 2:**
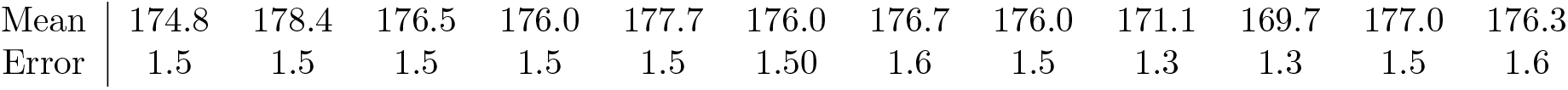
Collection of Jackknife bins Mean and Errors. We can see there is little statistical difference between the bins.

### 3.2 Stationarity Across Time of Day

Next, we consider stationarity across time of day, see Fig. 3 Left. In practice, this is what endocrinologists focus most of their attention on. As we can see the average blood sugar has varying steadiness depending on time of day. When the patient is awake the fluctuations in the BG shrink considerably and the mean blood sugar tends to be significantly lower than the A1C. This shows us that this particular patient’s CGM time series is not *stationary* across time of day. An important take away from this introduction is that there is NO guarantee that any CGM time series is in fact stationary. The stationary property is fundamental to time series analysis and it is an implicit assumption made by all previous studies [1, 2, 3, 4, 5]. If these authors followed LSTM guidelines, which calls for centering and rescaling the data, they unwittingly assumed stationarity across time of day as well. The centering and rescaling was likely done using complete time series average, ⟨*y*(*t*)⟩_all_, and the complete time series standard deviation. However, as we see in Fig. C Left, this assumption is only an approximation of the truth. I am hopeful however that, given ⟨*y*(*t*)⟩_days_ predominantly agrees ⟨*y*(*t*)⟩_all_ the error is likely small. As a general rule these results suggest that each patient’s data needs to be studied independently and recentered about the running mean ⟨*y*(*t*)⟩_days_ and NOT ⟨*y*(*t*)⟩_all_ to establish stationarity.

**Figure 3:**
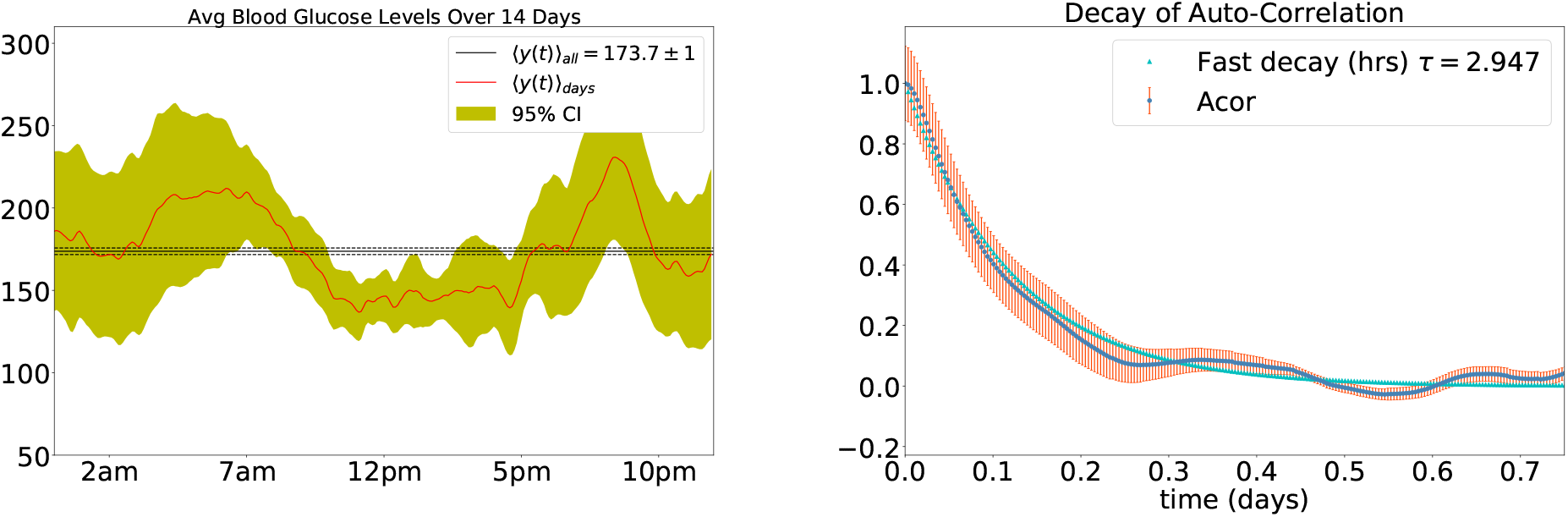
Left: Plot of patient’s 14 day average blood sugars, this plot demonstrates stationarity of the time series. Right: Plot of exponential decay of the auto-correlation (ACor) function, *τ*_*int*_ *≈* 2*τ*

## 4 Autocorrelation of CGM Time Series

From this CGM time series, we are able characterize the autocorrelation (**Acor**) in the data. The Acor decay rate *τ* = 2.883 hrs is presented in Fig.3. In Ref. [6], 2 decay times (*≈* 6 hrs) of an auto-correlated series is roughly the integrated Acor time *τ*_*int*_ (*i*.*e. τ*_*int*_ *≈*2*τ*). *τ*_*int*_ gives an estimate of how long it takes the time series to become decorrelated (*i*.*e*. two samples to become independent). Thus, this patient’s BG measurements are independent of what their BG was 6 hrs ago. Does this match expectation? First, consider the insulin. The patient in these trials was using a short acting insulin. Insulin’s activation curve demonstrates it is no longer effecting the patient after 2 decay times (*≈*6 hrs) Furthermore, in this study [7] it was found that postprandial spikes stabilizes in healthy patients after 6 hours. Since neither the food nor the insulin you took will be influencing you after 6 hrs, then the calculated decay rate of *≈* 3 hrs matches these physiological expectations.

When I attempt to fit the later portions of the ACor function with an oscillation, it is a very poor fit (Fig.4). In time series analysis periodic behavior is called *seasonality*, seasonality violates stationarity. So you test for it and remove it to ensure that the time series is stationary. We see that there is no clear periodicity in this diabetic’s CGM profile. In this study, subtracting off ⟨*y*(*t*)⟩_days_ from the time series shifts the center of the Acor function’s oscillating tail toward zero. The Fourier transform of the CGM time series is not shown here. The frequency domain is washed out in noise. This matches expectations that the body is a complex construction of interconnected systems. In fact, the T1D metabolic simulator [8] uses a set of 16 non-linear differential equations to create its FDA approved metabolic simulations. In conclusion, together the decay rate and non-periodic nature of the Acor show:

**Figure 4:**
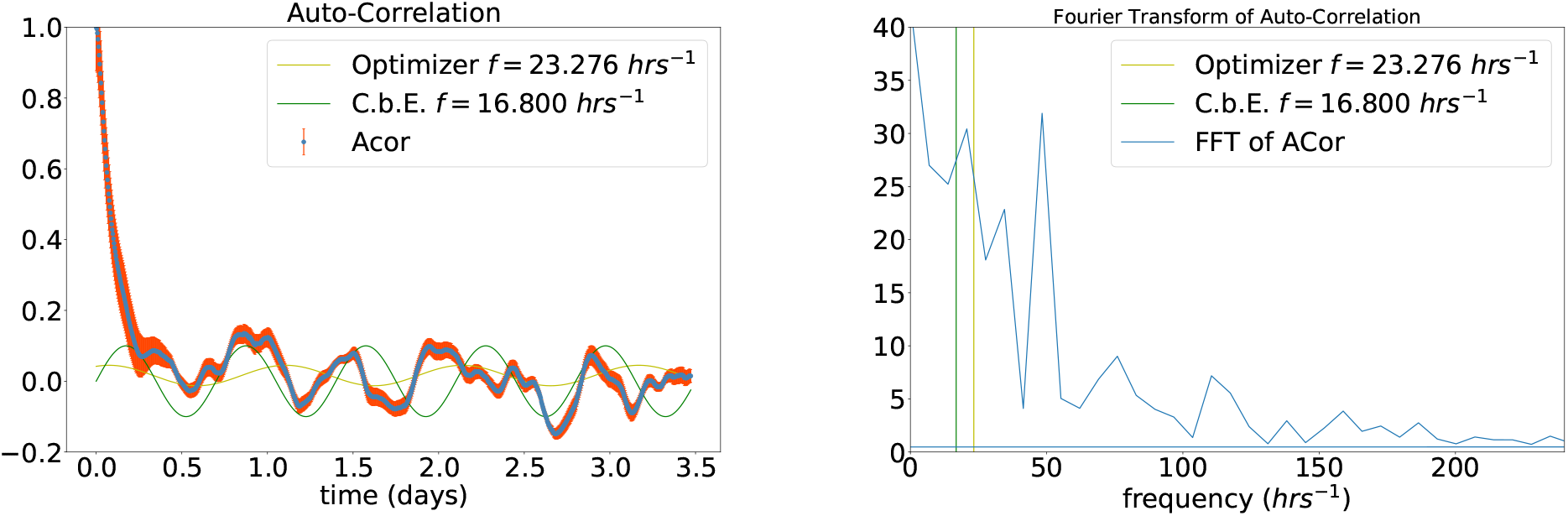
Left: Plot of oscillatory portion of auto-correlation (ACor). Yellow curve: Amplitude and Frequency both estimated by non-linear optimizer. Green curve: Amplitude and frequency estimated using “Chi by the eye” (C.b.E) technique. Right: Plot of the Fourier transform of the ACor function with the Sinusoidal frequencies plotted. We include plots of sinusoidal fits to demonstrate the non-periodicity in the oscillating tail of the Acor function.

1. Only the last 6 hours (2*τ*) could ever be useful for predicting the future.
2. Using regression to make predictions greater than 3 hours (*τ*) will have a great deal of uncertainty.
3. Depending on the patient’s physiology there could be different autocorrelations in their CGM data. As a result, pretraining an LSTM on another patient’s CGM, as in [3], is like to have had little effect.

## 5 Gaussian Process Regression

Gaussian process regression (**GPR**) is a kernel based (non-linear) regression methods. Its hyper-parameters can tuned to be either more or less sensitive to local versus global structure of the data. In practice, this tuning dampens or enhances steeper curvatures in the fit. Spuriously steep curvatures in the time series can be caused by noise. The faster the sampling rate the more prone the fit is to introduce these spurious oscillations. We normally tune the kernel parameters to make fits more robust to this noise, see Fig. 5 Left. By using the proper BG sampling rate and hyperparameters we can resolve both local and global structure. Fortunately, the GPR regression on the full CGM data resolves the peaks and avoids overfitting the data.

**Figure 5:**
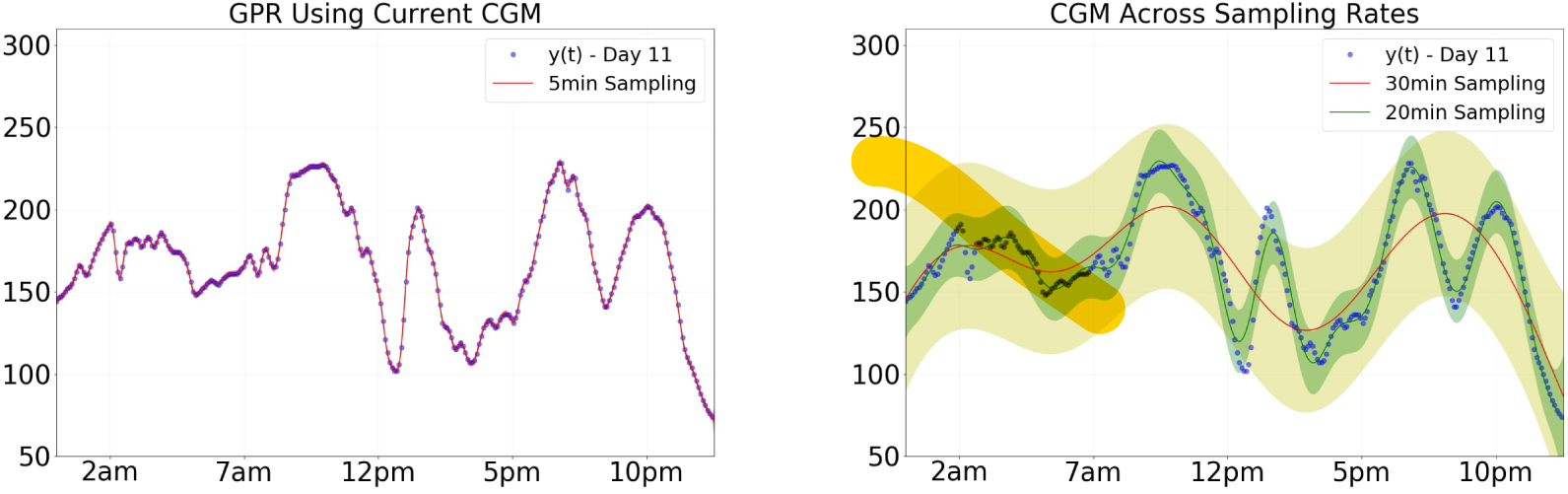
Initial tests of Gaussian Process Regression and their 95% confidence intervals across sampling rates. Left: GPR trained on full CGM data set, RMSE 3.03. Right: GPR trained with measurements removed from the full data set to decrease the sample rate to either 20 or 30 minutes apart. The RMSE values are 6.8 and 13.6 respectively. I conclude from the preserved structures and minimal RMSE that modern CGM’s have a sufficient sampling rate for time series analysis.

Next, I rarefy the sampling rate to study the CGM temporal resolution of the time series. Eventually, the sampling is decreased to point where the the global structure can no longer be resolved. This is known as aliasing, where two substantially different fits can reproduce the samples. ^2^ We search for this sampling threshold in Fig. 5 Right. In between 20 and 30 minute sampling rates, GPR loses its ability to resolve the fluctuations in the CGM. With a 30 minute sampling rate the GPR cannot resolve postprandial spikes[9, 7], somogyi effect [10], or dawn phenomena[11].

In conclusion, only the modern CGM wit sampling rates of 10 minutes or less can resolve a patient’s physiological responses. In this manner the current resolution is fine enough for the neural net to learn a model which can account for these peaks. This bodes well for the efficacy of an LSTM regression.

## 6 Characterizing LSTM predictions

In particular, we will study the LSTMs prediction when only CGM data is available (See Fig. 6). This matches the approach taken in [3]. This LSTM is based on Brownlee’s [12]. The network has a visible layer with 3 inputs, the most recent measurement and the previous 2 measurements. A hidden layer with 15 LSTM blocks with sigmoid activation functions. Finally, a dense output layer with tanh activation functions which makes the prediction. We use the first 14 days of the timeseries *≈* 4000 CGM measurements with an 80:20 training:testing split are used to train the network over 20 epochs, with a batch size of 10. Note, we display the CGM data at time *t* and the prediction at time *t* for time *t* + *t*_*p*_. As discussed in App. B lag time in the LSTM is expected. **I have soaked up the lag via a** *t* + *t*_*p*_ **interval shift for both linear and LSTM extrapolations**. I have shifted the prediction curve to demonstrate the LSTM’s feature preserving capacity.

**Figure 6:**
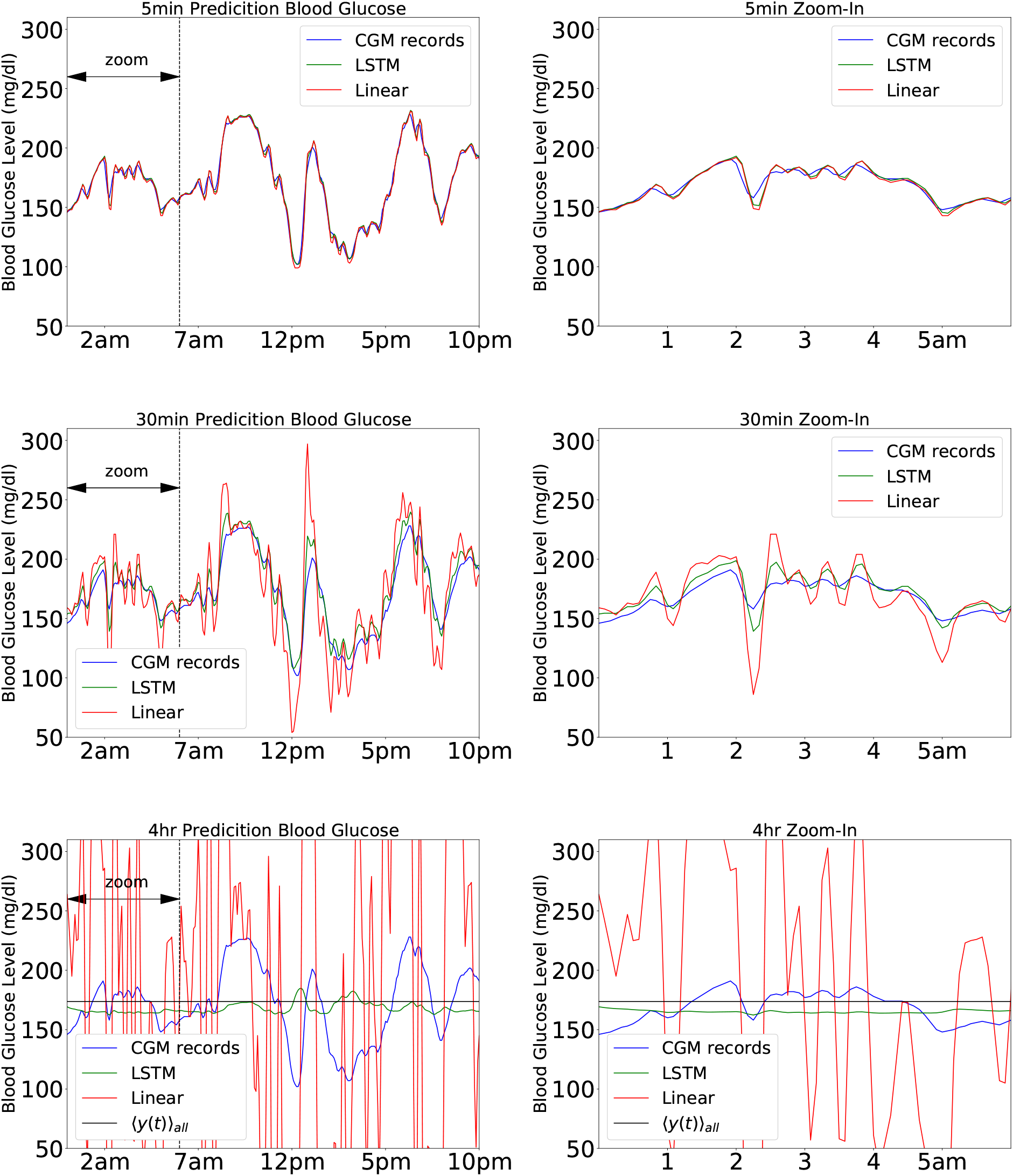
Comparison of LSTM neural network predictions to Linear extrapolation. **Top:** *t*_*p*_ = 5min prediction interval, LSTM Training RMSE 4.37, LSTM Testing RMSE 5.64. Linear RMSE 5.80. **Middle:** *t*_*p*_ = 30 min prediction interval, LSTM training RMSE 25.91, LSTM testing RMSE 22.15, Linear RMSE 32.53. **Bottom:** *t*_*p*_ = 4hr prediction interval: LSTM training RMSE 63.26, LSTM testing RMSE 42.27, Linear RMSE 281.42 **LSTM RMSE values are comprable to those quoted in [3]**. Left: Time interval centered on the Testing-Training domain split. Right: Time interval zoomed in on first six hours of the day.

This may make my prediction results seem better than they are. This is not my intent, by comparison the RMSE scores in Fig. 6 are on par with those in [3].

First, the flawless predictions at a 5 minute prediction interval are expected. Given the discussion in App.B, it takes longer than 5 minutes for physiological response to occur. **Thus, we expect that to predict 5 minutes into the future the LSTM does not have to learn any complicated non-linear functions which resemble those of physiological response**. Instead, a simple linear estimator would suffice. As we see in Fig.6, this is exactly the case.

Second, when the prediction interval is greater than the decay rate (*t*_*p*_ = 4*hrs > τ*), we see that the LSTM is guessing approximately the first moment of the stochastic process. Furthermore, the LSTM testing RMSE is 42 which is approximately the standard deviation of the testing data 40.5. For comparison, I have plotted the patients average blood sugar, ⟨*y*(*t*) ⟩_all_ alongside the LSTM predictions. The prediction interval is so large the data used to guess is uncorrelated (*i*.*e*. independent) to the predicted value. Meaning, the LSTM’s cost function converges to the ordinary least squares criteria for IID random variables. Even though the mean is the best linear unbiased estimator (**BLUE**), in this case, it is not a useful prediction. Thus, the decorrelation time, *τ*_*int*_, sets the upper bound on the prediction interval. Note that, even though the LSTM is failing at this extreme condition, its failure is mitigated with comparison to the linear approximation. As can be seen, simple linear extrapolation yields abhorrent results. The RMSE value of the linear extralopation, 281 is greater than the average BG concentration 173. However, Unlike the linear approximation methods, the LSTM is a smart algorithm; it has training and memory of its training. As argued, the LSTM has learns to guess ⟨*y*(*t*)⟩_all_, once it is asked to surpass the limits of predictability.

Third, the crucial plot for demonstrating the success of this LSTM is the middle one. At this prediction interval, the linear approximation breaks down yielding 150% greater RMSE then the LSTM testing. To elucidate this, focus on how the LSTM preserves the small peaks in the data while the linear approximation exacerbates them. The LSTM differs from linear extrapolations, in that the LSTM preserves the peaks as it is approaches the limit of its autocorrelated predictions. It intuitively follows from a first order Taylor approximation (*i*.*e*. linear estimate) of a function. The Taylor expansion has error on the same order as the square of the distance from the mean. Therefore the RMSE value of the linear extrapolation is roughly the same number as the prediction interval (*i*.*e. t*_*p*_ = 5 min and RMSE = 5.8, 30min - 32.53, 240min - 281.4). **Since the LSTM RMSE does not scale linearly with** *t*_*p*_ **it must be learning some non-linear model to predict patient BG concentration that gives it an edge over it’s linear competitor**. This prevents the closed loop from spiraling itself out of control at the prediction interval necessary for a closed loop to exist.

In summary, these numerical experiments show that the LSTM converges to a linear extrapolation in the limit of small prediction intervals. Furthermore, tests show that the LSTM prediction converges to ⟨*y*(*t*)⟩_all_ in the limit of large prediction intervals. There is a sweet spot around 30 minutes where the LSTM produces substantive predictions which outperform the linear extrapolation. Understanding these bounds has important implications in BG predictions.

## 7 Conclusions

Collectively, the complications from T1D reduce a patient’s life expectancy by 11 years for men and 13 years for women[13]. A preemptive response to blood sugar fluctuation can stave off the stock-piling of complications and extend the life-time of the diabetic. The ability to foretell BG has its greatest impact in type 1 diabetic (**T1D**) treatment plans. Early studies, like ([14, 15]), set the stage for the closed loop systems, which rely on on BG prediction techniques. The goal of the project is to characterize scenarios in which the LSTM will have trouble predicting and therefore managing blood sugar in a closed-loop insulin delivery system. As these neural network based approaches seep into biomedical treatment companies it will be important to understand the limitations of the method. Furthermore, it will be important to know how to implement the AIs. (*i*.*e*. how much and what kind of data to use to train it). This work has pinned down a few of those details. ^3^

1. In order for an AI to find BG prediction tractable it has to have a glucose monitor with a sufficient sampling rate. Modern CGM have an acceptable sampling to facilitate time series analysis.
2. Each patient’s BG time series is an ever changing non-stationary time series. There is an ever-advancing stationary window which limits how much data can be used to train the AI. The AI will need to be periodically retrained to keep up with this advancing window. Furthermore, stationarity across time of day is not guaranteed. So when training an LSTM centering and scaling needs to needs to be done not with the total sample average ⟨*y*(*t*)⟩_all_ and standard deviation, but with an time of day average ⟨*y*(*t*)⟩_all_ and its corresponding standard deviation.
3. Each patient’s BG levels have an inherent autocorrelation decay rate, this autocorrelation function will indicate any implicit periodic behavior and prediction interval *t*_*p*_ for regression techniques.
4. Each patient has a different physiological model, which at small prediction intervals can be approximated linearly. An LSTM learns non-linear models whose performance will vary from patient to patient depending on the autocorrelation inherent in a patient’s BG measurements. Meaningful predictions are not produced when the prediction interval exceeds the autocorrelation decay time, *τ*_decay_.

## A Introduction to Diabetes

### Pathology and Epidemiology

Type 1 Diabetes (T1D) is classified as a pancreatic disease and studied in endocrinology. In particular it is a medical condition in which the Beta cells in the pancreas fail to produce insulin. In fact, the beta cells of the pancreas are destroyed by the patient’s own auto-immune system. Typically T1D is developed after a sickness in which the over-active auto-immune system identifies the body’s own beta cells as targets. Thus, T1D is confirmed by the presence of autoantibodies, which are antibodies that target an individual’s own proteins. Empirically, the disease is more likely to affect families with diagnoses than without, so it is in someway hereditary. However, the underlying mechanism triggering the immune systems attack is yet unknown. Worldwide, the incidence of type 1 diabetes in children aged 14 years and younger has risen by 3% a year since the 1980’s. The current prevalence of the condition in the U.S. is roughly 1 in 300 by 18 years of age[16].

### Effects on Cellular Metabolism

T1D has a broad range of interconnected repercussions that happen in response to beta cell destruction. The beta cells create, store, and release insulin via the pancreas. Insulin is the main anabolic hormone of the body. As such, it regulates the metabolism of carbohydrates, fats and protein. It does so by promoting the absorption of glucose (derived as an end product of simple and complex carbohydrates) from the blood stream into the liver, fat cells, and skeletal muscle cells [17]. After absorbtion the glucose is converted into pyruvate^4^. The free energy released during this metabolic pathway is used to create ATP^5^. ATP is an adenine nucleotide bound to three phosphates, when your cells need energy they breaks these bounds. ATP is the body’s energy currency, like gas for a car. Insulin is therefore a lynchpin in your bodies metabolism process. Without insulin you body cannot remove the glucose from the blood stream. this has two important impacts ([18]).

1. The glucose stays attached to its carrier the red blood cell. Sugar crystals have sharp features, likewise, glucose’s chemical structure is incredibly sharp. The red blood cells carrying an over abundance of glucose will actually lacerate and damage the thinner capillaries of the body. The blood gets thick like honey is and puts increased work load on the heart to pump it.
2. The cells do not have access to the glucose needed for energy. To get the energy your body needs, the liver commits catabolism, it is like self cannabolism. In particular, liver begins the process of beta-oxidation. Where it breaks down macro-structures like fat and muscles to get monomers. Which it then breaks down in simple waste products classified as ketones (acetoacetate and *β*-hydroxybutyrate). The *β*-hydroxybutyrate can serve as a substitute energy source in the absence of insulin mediated glucose delivery.

The second impact is the same thing that happens when the body enters starvation. It is quite strange, the cells are starving standing next to a river full of food. However, because there’s no insulin they cannot access what they need.

### Physiological Symptoms

In the absence of insulin mediated glucose delivery the ketone byproducts of catabolism lead to an acidification of the blood stream. This acidification is aptly named Diabetic Ketoacidosis (**DKA**) [18]. It is a potentially fatal physiological state in which the patient experience vomiting, abdominal pain, deep gasping, increased urination, weakness, confusion, loss of consciousness, or comatose.

Insulin extraction and creation made it first break through in the early 1920’s being extracted from dogs. The first insulin shot was delivered in 1921. Over the next 70 years large scale insulin production became possible [19]. With the availability of insulin, T1D’s not only experience high blood glucose levels known as hyperglycemia, but also hypoglycemia (low BG levels) resulting from an insulin overdose. The symptoms of hypoglycemia are also potentially fatal irregular heart rhythm, fatigue, pale skin, shakiness, anxiety, sweating, hunger, irritability, confusion, blurred vision, seizures, loss of consciousness and diabetic shock.

Modern medicine has mitigated the symptoms which immediately lead to fatality. Thus, diabetics enjoy a longer life expectancy. However, diabetes is not yet a solved problem. There are long term complications that arise from diabetes. Such complications include: degradation of vision, kidney and liver failure, dementia, and amputation. All such complication occur after many years of intermittently high or low BGs.

## B Glucose Monitoring

Over the years glucose monitoring (GM) has evolved, in the early 20th century the sweetness of urine could indicate what the patient’s BG was hours before hand. In the late 50’s and early 60’s the first biosensor was developed which could determine the glycemia index from a sample of their blood. Then personal GM’s were marketed in the 80’s. In the 2010’s the continuous glucose monitor (**CGM**) came to the market. It is a biosensor that is attached to the patient and gives a constant readout of their BG[20]. With access to both insulin and blood glucose monitors, patients can fill-in the role of his or her pancreas with pen & paper calculations and medication. To stay alive, the modern diabetic is saddled with doing their pancreas’s job. The job is a constant balancing act, with insulin and exercise together forcing your blood glucose levels down and carbohydrates pushing the BG back up. However there is an input lag on insulin, food, and glucose monitoring. The producer of the drug Humolog, an artificial insulin, lists the specifications of the drug. It takes about 15 minutes to be absorbed and activated and has a 2 hour peak window in which it affects the body[21]. Additionally, after consuming food it takes at least 15 minutes for noticeable effects to occur in the blood stream. This lag estimate is further complicated by the type of food. The food’s glycemic index varies roughly inversely with the time to take effect[9]. Lastly, CGM systems measure glucose in interstitial fluid but not in blood. Rapid changes in one compartment are not accompanied by similar changes in the other, but follow with some delay. A study[22] shows that this lag is about 10 minutes. Combining these all together, after a meal we expect up to 30 minutes to pass till an effect is observed by a glucose monitor. The effect will last for about 2 hours. This lends itself to the timeseries provided by a CGM (whose average sample rate is 5 minutes) [23].

## C Introduction to Time Series Analysis

Practitioners of time series analysis need to determine the mean and the standard deviation of the mean (*i*.*e*. error) of various quantities from a stochastic process.

A *stochastic process* is a collection of random samples arranged in a sequence. As an example, a collection of dice roles indexed by the order in which they were rolled would count. When we sample data from a population we typically assume that the samples are independent and identically distributed (**IID**). However, this need not be the case. For example, your body weight which varies across your life and inside of any given 24 hour window. So if I took your weight 10 times in 5 minutes those would be IID samples. If I took your weight once a month for 5 years, and indexed over time. Then each samples would neither be independent of the next nor identically distributed as the next.

A *stationary* time series is one for which mean, variance, and autocorrelation do not change over observation time. Meaning all samples in the time series are identically distributed, but not necessarily independent. Mathematically, this is defined as:

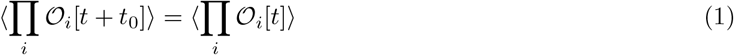

Meaning that the patient’s average blood sugar at 8pm do not vary significantly from their average blood sugar at 8am. A stationary time series is needed to be able to use all time series data to make predictions. Now that issue of identically distributed samples (*i*.*e*. stationarity) has been addressed let’s look at the condition of independent samples.

If two samples are correlated by definition they are not independent. *Auto-correlation* is the correlation between a measurement at time *t*_0_ with a measurement at time *t*_0_ + *t*. This is defined for a quantity *𝒪* in Ref. [6] as:

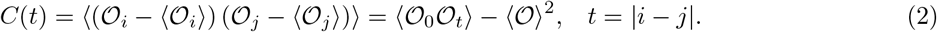

Where ⟨*𝒪*⟩_*i*_ is the mean value of the quantity *𝒪* at time *t*_*i*_. The normalized autocorrelation function is then

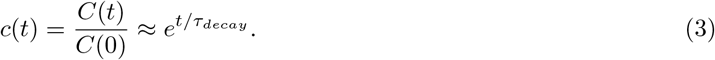

*c*(*t*) says that *𝒪* at time *t*_*i*_ is perfectly correlated to itself (*i*.*e. c*(0) = 1) as we would expect. As *t* increases, we expect the correlation to exponentially decay by Ref. [6]. Saying that the data is by definition equivalent to saying it is not independent. To account for this lack of independence, the variance of the mean (errorbars) need to be larger than the IID error by a factor called *the integrated autocorrelation time*:

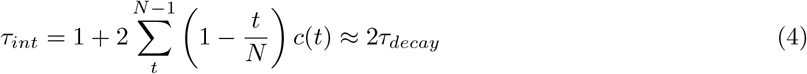

Alternatively, we could sample with time separation equal to *τ*_*int*_, at this interval the autocorrelation has decay sufficiently to be negligible. Hence we get away with assuming the samples are independent. Introductory statistics (IID samples) is recovered from time series analysis in the limit that we sample a *stationary* time series at intervals of *τ*_*int*_.

In the case of diabetes, blood glucose tests which are separated by 15 minutes are more likely to be the same than blood sugar tests separated by 24 hours. This is because your body’s next blood glucose measurement depends on its current blood glucose concentration. As a result, we expect the BG time series to have auto-correlation. Furthermore, the prediction of the next sample in a time series requires autocorrelation. To see this, consider a large prediction interval (*t*_*p*_ *> τ*_*int*_) then the best possible prediction value is the running mean of the data. The AI is attempting to predict the value of a decorrelated, IID, sample. Right off the bat, introductory statistics tells us that the best linear unbiased estimator for IID samples is the mean of the data. Thus, we need the data to be correlated so that we can make predictions other than the running meaning. As is the case with trivial machine learning (linear regression), the stronger the correlation between points the more accurate the prediction of the next point will be.

## D Regression

### D.1 Linear Regression

In typical regression, we consider some collection of centered measurements *x* = (*x*_1_, …, *x*_*n*_) such that *x*_*i*_ ∈ 𝒳 and 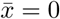. The goal is to predict some quantities *f* * from a possible measurements *x*\*. Assume that a function *f* such that *f* : 𝒳 → 𝒴 and that that function is linear, ⟨*y*_*i*_ = *f* (*x*_*i*_) + *e*_*i*_ = *βx*_*i*_ + *e*_*i*_. Where the error is assumed to be white noise є_*i*_ *∼* (𝒩, *σ*^∈^). Then a reasonable notion of error to judge the quality of a prediction is the least squares or squared error (**SE**) criteria,

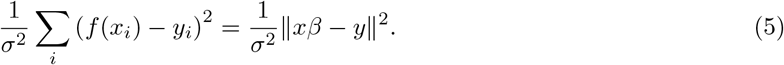

SE criteria yields a likelihood function. This is a probability distribution which describes the likelihood that any particular value of *β* is the best beta.

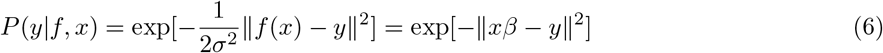

Low and behold this a Gaussian, so the most probable value is the value which maximizes the SE criteria. Mathematically, the Gaussian critical point is the best estimate of the linear coefficient *β*.

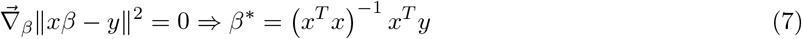

We predict values 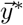 via the coefficient

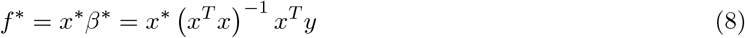

The estimate of *β* can be generalized via Bayes’ Theorem. Including a *a priori* estimate of the variance in our *β* parameters *P* (*β*), such that *β ∼* 𝒩 (0, *I*) Together this yields the Bayesian *a posteriori* estimate.

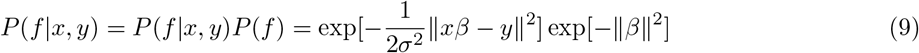

Which is maximized at *β*\* = (*x*^*T*^ *x* + *σ*^2^*I* − 1 *x*^*T*^ ⟨*y* yielding the predicted values

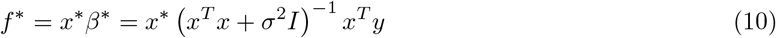

### D.2 Gaussian Process Regression

Gaussian Process regression (**GPR**) is different in its assumptions about the vector [*f* (*x*) = *f* (*x*_1_) … *f* (*x*_*n*_)]. Rather than assuming *f* (*x*) is a linear function. It assumes that *f* (*x*) are samples from a Gaussian process. This means that value *f* (*x*_*i*_) the *i*^*th*^ coordinate of a random sample from an *n*-dimensional Gaussian Eq. 11. The advantage of this assumption compared to assuming *f* (*x*) is a linear function is that: **GPR has the ability to create non-linear curves**.

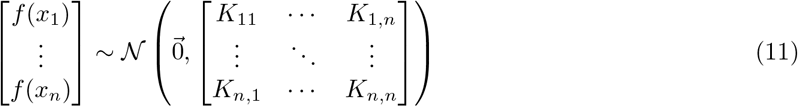

Note that 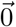 would be replaced by the mean function, E(*x*) if the data was not centered, (*i*.*e*. 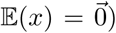). *K*_*i,j*_ = *K*(*x*_*i*_, *x*_*j*_) is the *covariance*. In practice, it can be any semi-positive definite kernel function. GPR is typically defined with *K*(*x*_*i*_, *x*_*j*_) = exp[− ∥*x*_*i*_ − *x*_*j*_∥ ^2^*/τ*]. Where *τ* is a hyperparameter that tunes how sensitive the regression is to the data’s overall global structure. By Schoenberg’s theorem, any exponentiated euclidean distance metric yields a semi-positive definite kernel. All regression is done to predict values *f* * = *f* (*x*\*) at given values *x*\*. The initial assumption that *f* is Gaussian distributed is of utmost importance in this process. The addition of two Gaussian variables *f* and *e* is itself a Gaussian variable *y*. As a result we can write the distribution of our joint vector *y, y*\* = *f, f* * + *e, e*\*

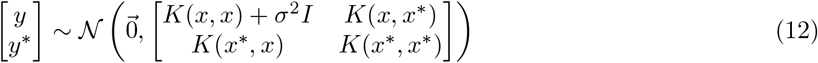

Fixing variables *y* to calculate the conditional probability of *y*\* we arrive at,

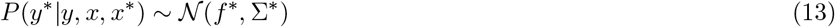

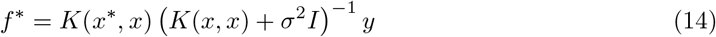

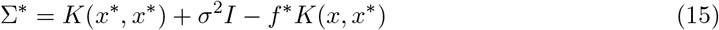

It is reassuring to notice that the predicted regression values in Eq. 14 resemble Eq. 10. Furthermore, each predicted value 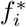 comes with error estimates 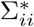. Using standard *Z*-scores we can calculate the 95% confidence interval from Eq. 15. Note that this appendix borrows heavily from Chuong B. Do’s course notes available at: http://cs229.stanford.edu/section/cs229-gp.pdf.

### D.3 Root Mean Squared Error

When judging the quality of a set of predictions, *y*\* against the actual measured values *y* it is standard to judge the quality of this set based on the RMSE. Which is defined as,

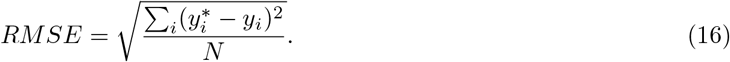

Eq. 16 has a similar form to the standard deviation of the mean. The average error will be zero, (*i*.*e*. we are equally likely to predict too high or too low). So this quantity indicates the spread of the error about its zero mean. The smaller the RMSE value the more tightly packed the errors are about zero. This is an important quantity for BG concentration because it gives a measure of how consistently accurate a blood glucose prediction system is.

## Data Availability

Transparency is of the utmost importance, data and analysis code are both available upon request. Please contact

## Footnotes

1 it has been noted that using physiological loss functions does not improve LSTM predictions [1]. This indicates that the LSTM is a form of personalized medicine, distinctly different from the typical corporation’s universal physiological models. The LSTM must be trained on each individual’s BG data rather than on a some collection of patients.

2 The relations between aliasing and sampling rates are characterized by the Nyquist sampling theorems.

3 Thanks is extended to Julie Libarkin for publication guidance, Michael Murillo for conversations regarding the methodology, and my old friend Sam Wittman for draft revisions.

4 This metabolic pathway is known as Gycolysis

5 If Gycolysis is done in an oxygen deficit environment, one of the byproducts is lactic acid. Lactic acid build up is what triggers muscle soreness after a work out.

